# Differential virulence potential of different clades of multidrug-resistant *Klebsiella pneumoniae* ST258

**DOI:** 10.64898/2026.03.28.26349612

**Authors:** Nathalie Chen, Brooke P. Dresden, Margaret Cassady, Marissa P. Griffith, Lora L. Pless, Lee H. Harrison, Ryan K. Shields, John F. Alcorn, Daria Van Tyne

## Abstract

*Klebsiella pneumoniae* (KP) isolates belonging to multi-locus sequence type 258 (ST258) are a frequent cause of hospital-associated outbreaks and display extensive multidrug resistance. The KP ST258 lineage consists of two genetically distinct clades, called Clade 1 and Clade 2. These two clades are genetically related to one another, but are historically distinguished by having different capsular polysaccharide types. While bacteria belonging to both clades are isolated from clinical infections, Clade 2 is isolated more frequently compared to Clade 1. To investigate drivers of this difference in clade prevalence, we collected 172 clinical KP ST258 isolates from patients at a single medical center. Clinical review showed that patients infected with Clade 2 isolates were more acutely ill than Clade 1-infected patients, despite having fewer comorbidities. We also found that Clade 2 isolates were more resistant to killing by human serum, despite binding more complement protein C3 than Clade 1 isolates. Additionally, mice infected with a Clade 2 isolate had increased bacterial dissemination from the lungs to the liver and spleen than mice infected with a Clade 1 isolate, and this dissemination required an intact capsule locus. Increased dissemination in mice was not due to differential serum killing, as mouse serum was unable to kill isolates of either clade, but dissemination was associated with decreased macrophage uptake of the Clade 2 isolate. Taken together, these data suggest that KP ST258 Clade 2 is more virulent than Clade 1, though the specific mechanisms at play appear to differ between mice and humans.

**IMPORTANCE:** KP ST258 is an epidemic lineage of multidrug-resistant gram-negative bacteria that has caused numerous outbreaks in hospitals around the world. The KP ST258 population is divided into two genetically related but distinct clades, which differ primarily in their capsule type. In this study, we found that patients infected with one of the KP ST258 clades were more acutely ill than patients infected with the other clade. We also observed clade-specific differences in killing by human serum and bacterial dissemination in a mouse model of pneumonia. Finally, we identified important limitations in the use of mouse models to study host defenses against multidrug-resistant KP infection. Overall, this work underscores the importance of capsule composition in KP ST258 virulence, identifies differences in the host response to KP infection between mice and humans, and highlights a potential role for complement-targeting immunotherapeutics in KP treatment.

## INTRODUCTION

*Klebsiella pneumoniae* (KP) is a gram-negative pathogen that frequently causes hospital-acquired infections (1, 2). KP has become increasingly multidrug-resistant, and carbapenem-resistant KP is considered a top priority pathogen by both the US Centers for Disease Control and Prevention and the World Health Organization (3, 4). The classical pathotype of KP is frequently multidrug resistant, however it is thought to cause infections primarily in immunocompromised patients. This is in contrast to the hypervirulent pathotype of KP, which is more susceptible to antibiotics but is capable of causing invasive infections in immunocompetent patients (5, 6). The possibility of convergence of these two pathotypes, resulting in multidrug-resistant isolates capable of invasive infections, is a looming global health threat (7, 8). A better understanding of classical KP lineages and how they cause infections can help identify new therapeutic targets effective against classical, hypervirulent, and convergent KP.

Sequence type 258 (ST258) is a prominent classical KP lineage that has caused numerous outbreaks of carbapenem-resistant infection around the globe (9–13). Although the extensive multidrug resistance of KP ST258 contributes to its dissemination, it is not the only KP lineage that carries carbapenem resistance and the mechanisms underlying its success as an epidemic lineage have yet to be fully elucidated. The KP ST258 population is divided into two closely related clades, called Clade 1 and Clade 2, that differ primarily in their capsular polysaccharide type (14). Capsule is a major virulence factor for KP, and over 160 different capsule types have been described (15). Different capsule types have been previously associated with different degrees of virulence in both *in vitro* experiments and *in vivo* in mouse models of infection (16–18). However, it is unknown whether differences in capsule type between the two KP ST258 clades impact the ability of each clade to cause disease.

Prior head-to-head comparisons of Clade 1 and Clade 2 KP ST258 isolates have been limited. Genomic comparisons of KP ST258 populations have found a strict association between clade and carbapenemase type (14, 19, 20). The two clades have also been shown to differ in their plasmid content, and prior studies found that Clade 1 isolates tend to have more antibiotic resistance genes than Clade 2 isolates (14, 19, 20). One prior study found that there was no difference in human serum survival between seven Clade 1 isolates and 13 Clade 2 isolates (21). Other studies have shown that a representative Clade 2 isolate induced more production of IL-1β, IL-6, IL-17A, IL-23 and TNFIZ from cultured human myeloid dendritic cells and caused increased differentiation of Th17 cells in cultures of human peripheral blood mononuclear cells than a representative Clade 1 isolate (22, 23). Several studies have also reported that Clade 2 is more common in isolate collections (24–26), suggesting that Clade 2 isolates may be more virulent than Clade 1 isolates. The existence of two genetically related clades that differ in their capsule type offers an opportunity to examine the role that KP capsule composition (rather than capsule abundance) plays in pathogen virulence. In this study, we systematically assessed the clinical features and *in vitro* phenotypes that distinguish Clade 1 from Clade 2 infections to evaluate if KP ST258 Clade 2 isolates are indeed more virulent than Clade 1 isolates.

## RESULTS

### KP ST258 Clade 2 isolates are associated with more severe infections than Clade 1 isolates

Between 2015 and 2024, 172 clinical isolates of carbapenem-resistant *K. pneumoniae* were collected from the University of Pittsburgh Medical Center (UPMC) and identified as belonging to ST258. Consistent with prior reports, over 70% of isolates were identified as belonging to ST258 Clade 2 (n=46 Clade 1 isolates, n=126 Clade 2 isolates) (**Fig 1A**). The distribution of isolate sources differed by clade, with Clade 1 isolates primarily collected from urine specimens and Clade 2 isolates primarily collected from respiratory specimens (**Fig 1B**, *P*<0.0001). To determine if there were differences in the types of patients infected with Clade 1 versus Clade 2 isolates, or if infection outcomes differed by clade, we reviewed the clinical records of all 172 patients. Patients infected with Clade 1 isolates did not differ in their age from patients infected with Clade 2 isolates, however they were more often female (63% versus 43%, *P*=0.0248) (**Table 1, Fig S1A**). We used the Charlson Comorbidity Index to assess each patient’s overall health at the time their isolate was cultured (27), and observed that patients infected with Clade 2 isolates had, on average, a lower Charlson Comorbidity Index than patients infected with Clade 1 isolates (4 [3-6] versus 5.5 [3-8.75], *P*=0.0297) (**Table 1, Fig S1B**). Despite having fewer comorbidities on average, patients with a Clade 2 infection were more acutely ill compared with Clade 1-infected patients, as measured by the Pitt Bacteremia Score (3 [1-5] versus 2 [0-3], *P*=0.0085) (**Table 1, Fig S1C**) (28, 29). Clade 2-infected patients also spent more cumulative days in the intensive care unit (ICU) than Clade 1-infected patients, suggesting that their infections were associated with more complex hospitalizations (**Table 1**, **Fig 1C**, *P=*0.0043). We also observed a trend towards a lower probability of 14-day survival among patients infected with Clade 2 isolates, however this difference was not statistically significant (**Fig 1D**, *P*=0.0757). Together these clinical measures suggest that Clade 2 isolates might be more frequently sampled because they are associated with more severe infections.

**Fig 1.**
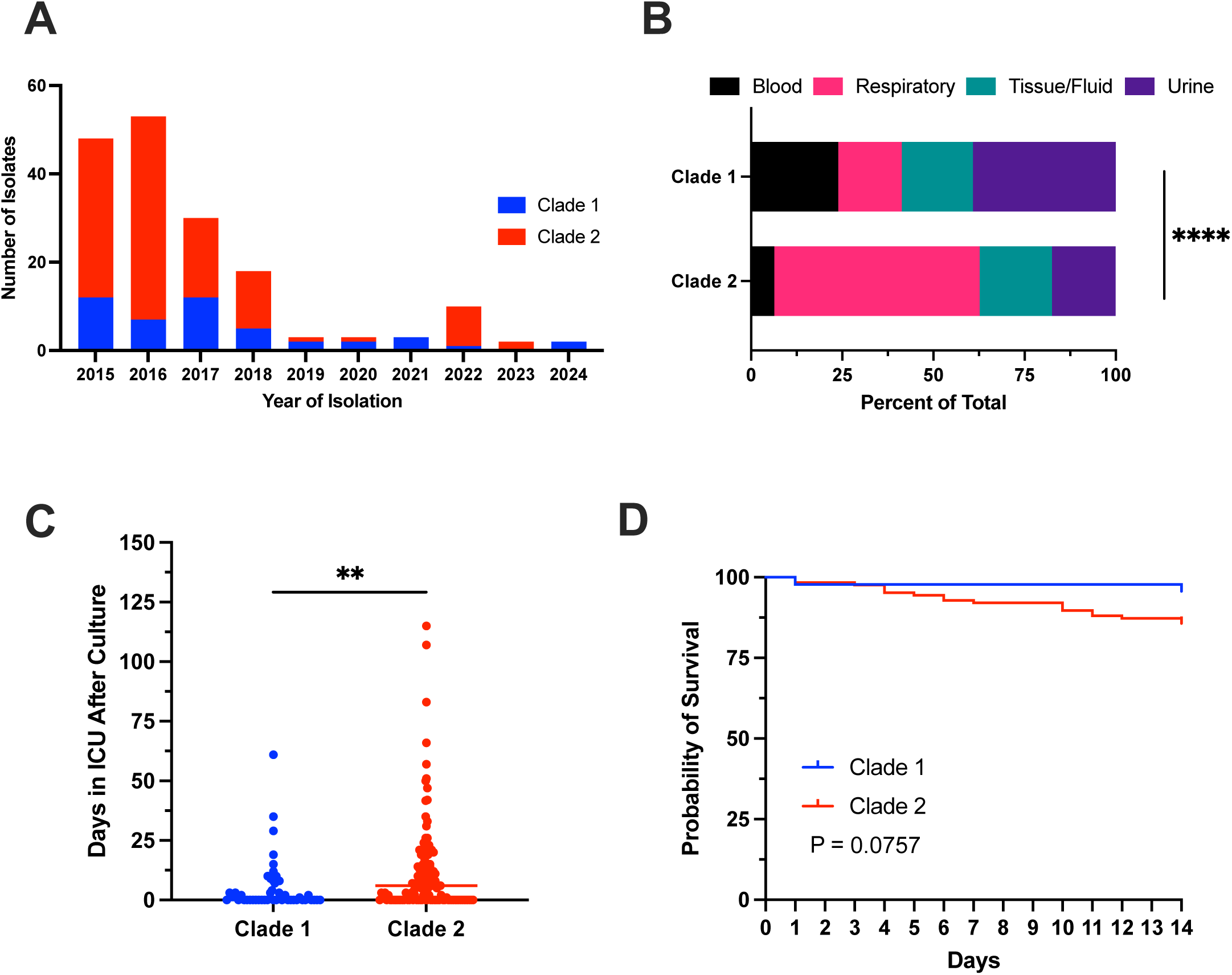
Clinical features of 172 patients infected with KP ST258. (A) 172 carbapenem-resistant KP ST258 isolates were collected from unique hospitalized patients between 2015 and 2024. (B) Source distribution of isolates in each clade. Blood specimens include those from blood cultures and central venous catheters. Respiratory specimens include those derived from sputum, bronchoalveolar lavage fluid, bronchial wash, sputum, and tracheal aspirate. Tissue/Fluid specimens include those derived from tissue, wounds, ulcers, pleural fluid, ascites fluid, and drains associated with bodily fluids. Urine specimens include those derived from urine cultures. Significance was assessed using Fisher’s exact test. P < 0.0001: ****. (C) Number of cumulative days spent in the intensive care unit (ICU) for each patient after their isolate was cultured. Significance was assessed by Mann-Whitney test. P < 0.01: ** (D) Survival of patients infected by Clade 1 or Clade 2 isolates for 14 days after the clinical isolate was obtained. P*-*value is calculated from a log-rank test.

**Table 1.** Patient Characteristics.

|  | **Clade 1**<br>(n=46) | **Clade 2**<br>(n=126) | **P-value** |
| --- | --- | --- | --- |
| Age (years) (median (range)) | 65.5 (31-91) | 64.0 (23-95) | 0.4219 * |
| Sex (% female) | 63.04 | 42.86 | 0.0248 ^ |
| Charlson Comorbidity Index (median (IQR)) | 5.5 (3-8.75) | 4 (3-6) | 0.0297 * |
| Pitt Bacteremia Score (median (IQR)) | 2 (0-3) | 3 (1-5) | 0.0085 * |
| Length of Hospitalization<br>(median number of days (IQR)) | 21.0 | 34.5 | 0.0315 * |
| Cumulative length of ICU stays after culture<br>(median number of days (IQR)) | 0 (0-6.25) | 6 (0-15) | 0.0043 * |

\* Mann-Whitney test
^ Fisher's Exact test

### Clade 2 isolates are more closely related and encode more siderophores than Clade 1 isolates

To understand why Clade 2 isolates are isolated more frequently and why they might be associated with more severe infections compared with Clade 1 isolates, we analyzed the available genomes of 132 KP ST258 isolates from study patients (n=25 Clade 1, n=108 Clade 2). A maximum likelihood phylogenetic tree showed the characteristic two-clade structure of ST258 (14) (**Fig S2**). Consistent with prior reports, all Clade 1 isolate genomes encoded the KL106 capsule locus, while all Clade 2 isolates encoded the KL107 locus. We purified the capsular polysaccharide from representative isolates of each clade (Clade 1: KLP00155, Clade 2: KLP00157) and performed carbohydrate composition analysis, which showed that the Clade 1 isolate had a predominantly glucose-based capsule whereas the Clade 2 isolate had a predominantly rhamnose-based capsule (**Table S1**), in agreement with previously published structures (30, 31). There was no significant difference between the genome sizes of Clade 1 and Clade 2 isolates (**Fig 2A**), however split kmer analysis showed that Clade 2 isolates were more genetically similar to each other, as measured by pairwise single nucleotide polymorphisms (SNPs) (**Fig S3A**, *P<*0.0001). Through average linkage clustering with a 10-SNP cutoff, we identified one putative transmission cluster containing two closely related Clade 1 isolates, and 12 clusters containing 36 Clade 2 isolates (**Table S2 and S4, Fig S2**), suggesting that Clade 2 isolates were more likely to be involved in putative transmission clusters compared with Clade 1 (**Fig 2B**, *P=*0.0128).

**Fig 2.**
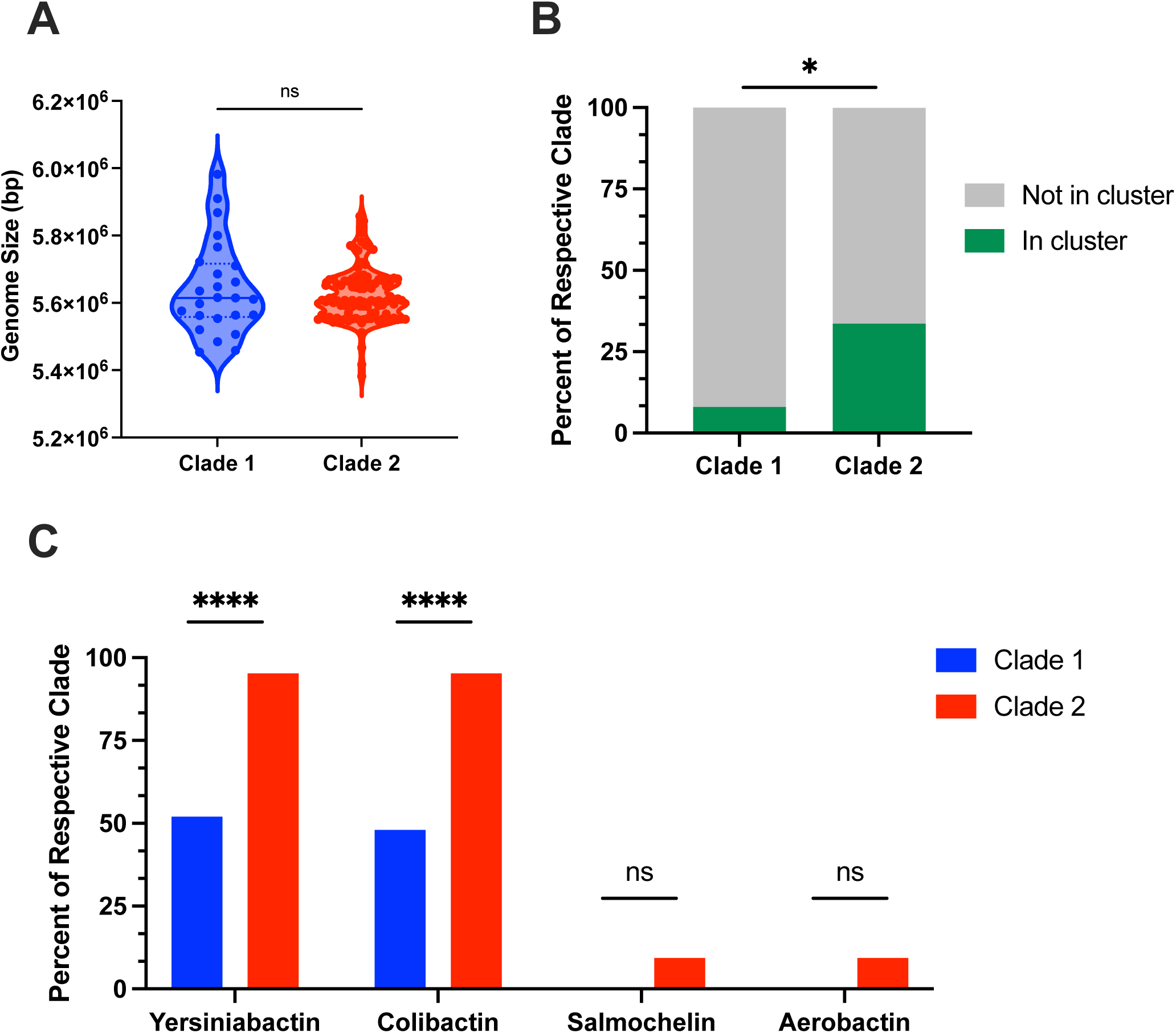
Clade 2 isolates are more closely related and encode more siderophores than Clade 1 isolates. 132 ST258 genomes (Clade 1 = 25, Clade 2 = 107) were assessed for: (A) Genome size using Kleborate; (B) Genetic relatedness using Split Kmer Analysis and average linkage clustering with a 10-SNP cutoff; (C) Siderophores in isolate genomes identified using Kleborate. Significance was assessed with Mann-Whitney test (A) and Fisher’s exact test (B and C). P ≤ 0.05: *, P ≤ 0.0001: ****.

While increased nosocomial transmission of Clade 2 isolates could explain why they are more frequently isolated from patients, it does not explain why they appear to be associated with worse clinical outcomes than Clade 1 isolates. To investigate potential determinants of this association, we assessed plasmids, antibiotic resistance genes, and known virulence factors that were differentially present in Clade 1 versus Clade 2 isolate genomes. Consistent with their greater genetic diversity, Clade 1 isolates encoded more plasmid replicons and more antibiotic resistance genes than Clade 2 isolates (**Fig S3B** *P<*0.0001; **S3C** *P=*0.0021). The difference in antibiotic resistance genes was driven by Clade 1 isolates more frequently encoding resistance to macrolides and phenicols (**Fig S3D**, both *P<*0.0001), however neither of these drug classes are first-line treatments for multidrug-resistant gram-negative infections such as those caused by KP ST258 (32). We next examined the differential presence of siderophores, which are well-characterized virulence factors for classical KP (2), and found that Clade 2 isolates more frequently encoded the siderophores yersiniabactin and colibactin (**Fig 2C**, both *P<*0.0001). Taken together, these data suggest that Clade 2 isolates might be more common among clinical collections of KP ST258 because they are more frequently transmitted in the hospital, and could be associated with sicker patients at least in part because they encode more siderophores than Clade 1 isolates.

### Clade 2 isolates demonstrate increased resistance to human serum killing despite increased complement activation

Although Clade 2 isolates more frequently encoded siderophores, half of Clade 1 isolates also encoded yersiniabactin and colibactin (**Fig 2C**). In addition to siderophores, capsule is also a critical virulence factor for KP (2, 33, 34). Because differing capsule types are a defining feature of the two clades of KP ST258, we wondered how capsule composition might impact the virulence of each clade. To compare the virulence of isolates in each clade in a translationally relevant *in vitro* assay, we quantified bacterial survival in human serum for 13 Clade 1 and 36 Clade 2 isolates. We found that Clade 2 isolates were more frequently able to resist serum-mediated killing compared with Clade 1 isolates (**Fig 3A**, *P*=0.026). To determine if this difference was due to differences in capsule composition, we tested if removal of the capsule locus eliminated clade-specific differences in serum killing. We did this by deleting the capsule locus and replacing it with a tetracycline resistance cassette in a representative Clade 1 and Clade 2 isolate studied previously (19), generating KLP00155Δcps and KLP00157Δcps strains. We verified that capsule knockout strains had minimal capsule production compared to their parent strains (**Fig S4A**), and also observed slight growth defects compared to the parent strains, especially for the Clade 1 KLP00155Δcps strain (**Fig S4B and C**). We then tested how capsule locus removal impacted serum survival, and found that both capsule knockout strains were equally highly susceptible to killing by human serum (**Fig 3B**). These data suggest that the observed difference in serum survival of the parent strains could be due to the different capsular polysaccharides that they produce.

**Fig 3.**
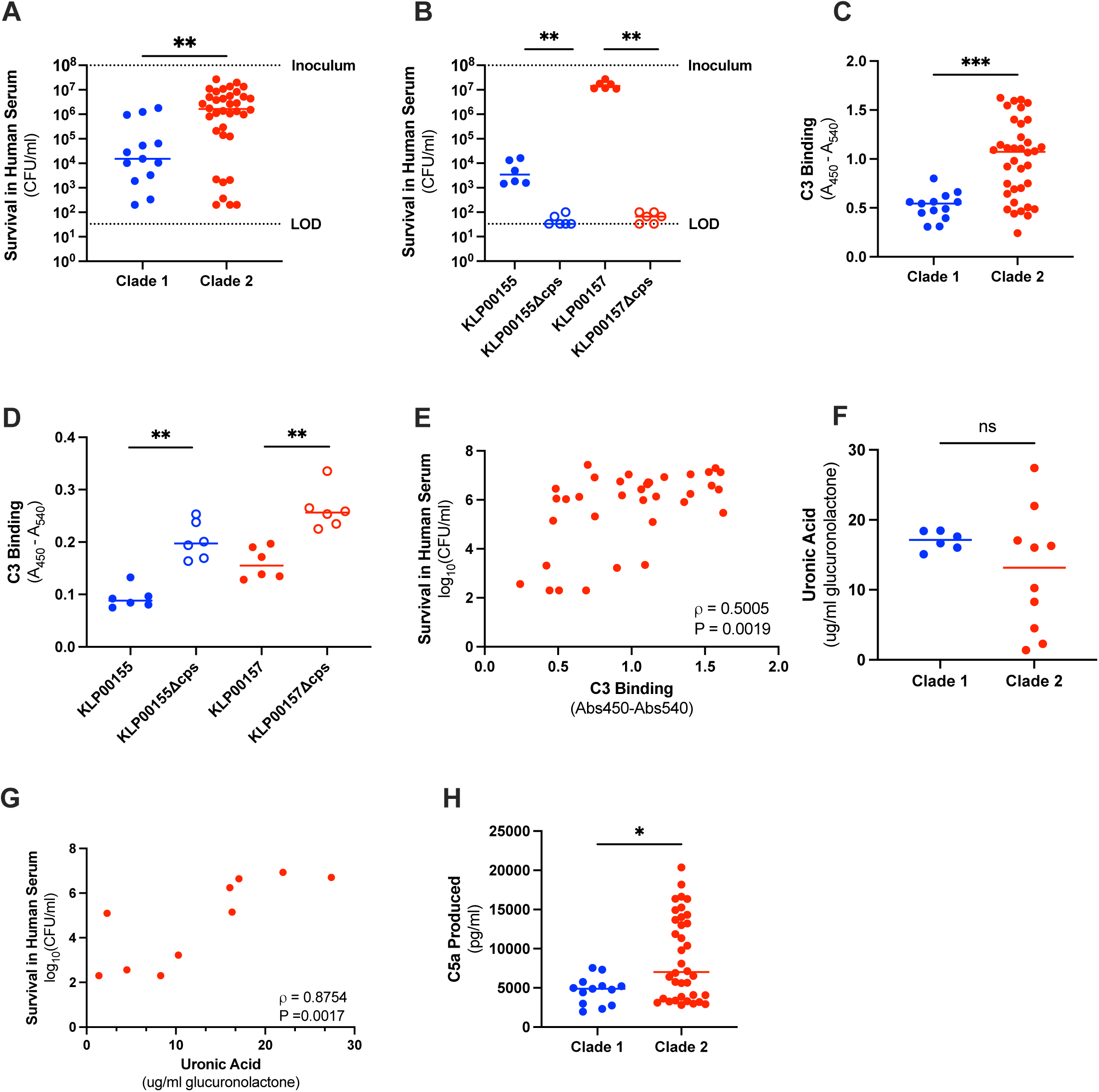
Clade 2 isolates are more resistant to killing by human serum than Clade 1 isolates, despite increased complement binding and activation. (A and B) Bacteria recovered after 30-minute exposure of 10^8^ CFU/mL to 85% human serum. Limit of detection (LOD) is indicated with a dashed line. (A) 13 Clade 1 isolates and 36 Clade 2 isolates were tested, each with at least two biological replicates. (B) 6 biological replicates per strain were tested. (C and D) C3 binding to bacteria was measured using an indirect ELISA. (C) 13 Clade 1 isolates and 36 Clade 2 isolates were tested, each with at least two biological replicates. (D) 6 biological replicates were tested per strain, each with at least 5 technical replicates. (E) Correlation between C3 binding and serum survival of Clade 2 isolates was assessed using Spearman correlation. (F) Uronic acid was quantified from a selection of Clade 1 (n=6) and Clade 2 (n=10) isolates with 3 biological replicates per isolate. Significance was assessed with Welch’s t-test. (G) Correlation between uronic acid quantity and serum survival of 10 Clade 2 isolates was assessed using Spearman correlation. (H) OD-normalized bacteria were reacted with 5% human serum in PBS for 30 minutes. C5a from the supernatant of the reaction was quantified using a sandwich ELISA. At least two biological replicates per isolate were performed. For panels A-D and H, P ≤ 0.05: *, P ≤ 0.01: **, P ≤ 0.001: *** by Mann-Whitney test.

Killing by human serum can be mediated by several different components, and it has been suggested that an important factor for KP killing is the complement system (21). To begin to investigate possible mediators of the differential susceptibility of Clade 1 and Clade 2 isolates to human serum, we assessed the binding of complement protein C3 to each isolate using indirect ELISA. We found that Clade 2 isolates displayed a wide range of C3 binding, but on average they bound more C3 than Clade 1 isolates (**Fig 3C**, *P=*0.0002). Capsule knockout strains each had increased C3 binding compared to their parent isolate (**Fig 3D**, both *P=*0.0022). While there was no significant correlation between C3 binding and survival in human serum among Clade 1 isolates (**Fig S5A**), we did observe a significant correlation among Clade 2 isolates whereby isolates that bound more C3 had increased serum survival (**Fig 3E**). We also measured capsule production in a subset of Clade 1 and Clade 2 isolates and found that capsule production was highly variable and significantly correlated with serum survival among Clade 2 isolates, but not Clade 1 isolates (**Fig 3F, G; Fig S5B**). Increased C3 binding by Clade 2 isolates led us to wonder if these isolates might also display increased downstream complement activation compared to Clade 1 isolates. Indeed, we found that incubation of Clade 2 isolates with human serum resulted in increased release of the anaphylatoxin C5a compared with Clade 1 isolates (**Fig 3H**, *P*=0.0155). Overall, these data indicate that KP ST258 Clade 2 isolates have increased resistance to killing by human serum that is likely mediated by their capsule. Additionally, serum resistance among Clade 2 isolates is correlated with increased C3 deposition and more C5a production, suggesting that these isolates employ a mechanism of serum resistance that does not rely on evasion of complement deposition (35).

### A Clade 2 isolate disseminates more than a Clade 1 isolate in a mouse model of pneumonia

To investigate whether the *in vitro* virulence differences we observed between Clade 1 and Clade 2 isolates translated to differences in *in vivo* virulence, we tested representative isolates in a mouse model of pneumonia. We first infected mice via oropharyngeal aspiration with 10^8^ CFU of representative Clade 1 (KLP00155) or Clade 2 (KLP00157) isolates. We found that KLP00157-infected mice died more quickly than KLP00155-infected mice (**Fig 4A**, *P=*0.0023), however the high inoculum resulted in rapid death of nearly all animals regardless of clade. To understand why the KLP00157-infected mice might have died more quickly, we next infected mice via oropharyngeal aspiration with a lower inoculum (10^7^ CFU) of either KLP00155 or KLP00157 and assessed lung injury, lung inflammation, and bacterial dissemination at 6- and 24-hours post-infection. To assess lung injury and inflammation, we measured protein content in bronchoalveolar lavage fluid (BALF) (**Fig S6A**), number of cells in BALF (**Fig S6B**), and cell counts in BALF of monocytes, polymorphonuclear cells, eosinophils, and lymphocytes (**Fig S6C**). At both timepoints, there was no significant difference between the KLP00155-infected and KLP00157-infected mice in all measures (**Fig S6A, B, C**). We also measured the expression of IL-1β, IFN-LJ, IL-6, TNF-IZ, CXCL1, CXCL5, IL-23A, and lipocalin 2 (LCN2) in lung tissue using qRT-PCR. Besides a modest increase in IL-6 expression at 24-hours post-infection in KLP00157-infected mice, there were no significant differences in lung cytokine profiles between KLP00155-infected and KLP00157-infected mice (**Fig S6D and E**). We did find, however, that KLP00157-infected mice had increased dissemination of bacteria from the lungs to the spleen at 6-hours post-infection, and increased dissemination to the spleen and liver at 24-hours post-infection, compared to KLP00155-infected mice (**Fig 4B, C**). To determine if this increased bacterial dissemination was a capsule-mediated process, we next infected mice with 10^7^ CFU of either KLP00157 or KLP00157Δcps, and found that deletion of the capsule locus led to increased clearance of bacteria from the lungs and greatly reduced dissemination to the spleen and liver at 24-hours post-infection (**Fig 4D**). These data suggest that Clade 2 might be more virulent in a pneumonia model of infection due to its increased ability to disseminate, and that the Clade 2 capsule likely plays an important role in dissemination.

**Fig 4.**
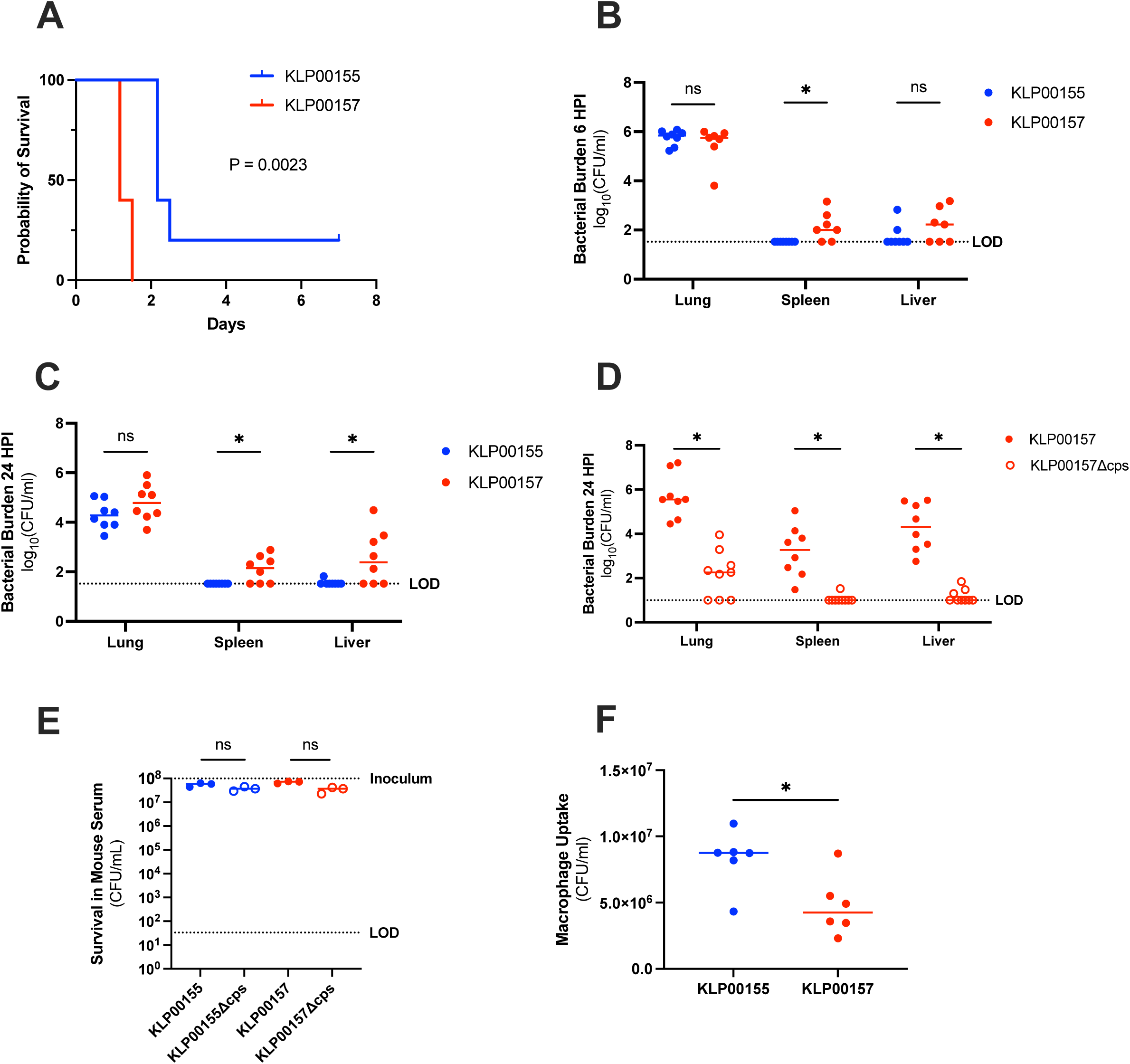
Clade 2-infected mice have increased bacterial dissemination. 8-10 week-old female C57BL/6J mice were infected via oropharyngeal aspiration. (A) Mice were infected with 10^8^ CFU of either KLP00155 (n=5) or KLP00157 (n=5). Survival was assessed using a log-rank test. (B and C) Mice were infected with 10^7^ CFU of either KLP00155 or KLP00157 and the upper right lung lobe, spleen, and liver were harvested for bacterial enumeration at (B) 6-hours (KLP00155 n=8 and KLP00157 n=7) or (C) 24-hours (KLP00155 and KLP00157 n=8) post infection. (D) Mice were infected with 10^7^ CFU of either KLP00157 or KLP00157Δcps and the upper right lung lobe, spleen, and liver were harvested for bacterial enumeration at 24-hours post-infection (KLP00157 n=8 and KLP00157 Δcps n=9). (E) Bacteria recovered after 30-minute exposure of 10^8^ CFU/mL bacteria to 85% mouse serum. Three biological replicates were performed per strain. (F) Immortalized mouse bone marrow-derived macrophages were incubated with live KLP00155 and KLP00157 for 2 hours (MOI=10) followed by an additional hour with gentamicin. The macrophages were then lysed and plated for CFU determination. Six biological replicates were performed with at least two technical replicates each. Limit of detection (LOD) is shown with a dashed line for each relevant panel. For panels B-F, P ≤ 0.05: * by Mann-Whitney test.

Because Clade 2 isolates were more resistant to being killed by human serum, we hypothesized that KLP00157 might disseminate more than KLP00155 in mice because it could resist serum-mediated killing, thereby conferring increased fitness in the bloodstream. When we repeated the *in vitro* serum killing assay using mouse serum, however, we found that both KLP00155 and KLP00157, as well as the capsule knockout strains, were entirely resistant to killing by mouse serum (**Fig 4E**). An alternative hypothesis to explain why KLP00157 disseminated more than KLP00155 could be that KLP00157 is less effectively cleared by tissue-resident immune cells, such as macrophages. We therefore assessed the degree to which KLP00155 and KLP00157 were taken up by murine bone marrow-derived macrophages, and found that there was more uptake of KLP00155 compared with KLP00157 (**Fig 4F**, *P*=0.026). Taken together, these data suggest that Clade 2 isolates might have increased virulence due to their ability to disseminate to other organs, but the mechanism(s) underlying this increased dissemination might be different between mice and humans.

## DISCUSSION

Carbapenem-resistant KP infections remain an important problem in healthcare settings. Although KP ST258 prevalence is decreasing (36, 37), the presence of two clades with different capsule types within ST258 offers a unique opportunity to gain insight into the clinical importance of capsule type in KP infection. While the capsule types of these two clades have been structurally characterized and shown to be quite different, the impact of these differences have yet to be elucidated. Prior studies investigating how capsule type impacts virulence independent of genetic background focused on *in vitro* phenotypes and murine infection models (16, 18, 38), but it remains to be seen if those findings extend to human infections. Further, prior studies directly comparing the two clades of KP ST258 utilized small numbers of representative isolates, hindering generalizability. In this study, we leveraged a large collection of clinical KP ST258 isolates to systematically measure the clinical impact of clade on pathogen virulence *in vivo* in human patients, *in vitro* in assays that reflect virulence in humans, and in a mouse model of infection. Our findings suggest that ST258 Clade 2 isolates are more virulent than Clade 1 isolates in both humans and mice, but the mechanisms underlying this difference appear to be distinct between these different mammalian hosts.

We identified important clinical differences between patients at our hospital infected with Clade 1 and Clade 2 isolates. In alignment with prior studies (24–26), we found that infections at our center were predominantly caused by Clade 2 isolates. We also found that, despite having lower comorbidity scores, patients with Clade 2 infections had higher Pitt Bacteremia Scores and spent more cumulative days in the ICU post-culture. We also identified a trend towards increased mortality among Clade 2-infected patients, again suggesting that Clade 2 isolates cause worse disease in susceptible hosts. Although this trend was not statistically significant, it does align with a prior study that found infection with *wzi*-154-encoding ST258 isolates (which are often ST258 Clade 2) was associated with increased mortality compared to non-*wzi*-154-carrying carbapenem-resistant KP, though the association was borderline significant likely due to small sample size (39). The increased virulence of Clade 2 isolates could help explain why the clade is more common than Clade 1 among clinical collections of KP ST258 isolates, as its increased virulence might make it more likely to be sampled from sick patients.

Through comparative genomic analysis and clustering, we found potential explanatory mechanisms for our clinical observations. We observed that Clade 2 isolates were more closely related to one another and more often resided in putative transmission clusters compared with Clade 1 isolates, which could contribute to why Clade 2 was more frequently isolated than Clade 1. In agreement with findings from a prior study at our center (19), we found that Clade 1 isolates had more plasmids and antibiotic resistance genes compared with Clade 2 isolates, however the resistance genes driving this difference encoded macrolide and phenicol resistance, neither of which would be used as first-line treatment for KP infections. We did, however, observe that nearly all Clade 2 isolates encoded the siderophores yersiniabactin and colibactin, while only about half of Clade 1 isolates did so. This might help explain why Clade 2 isolates were more frequently isolated from respiratory specimens, as yersiniabactin in particular has been shown to help KP survive in the respiratory tract (40).

*In vitro* experiments revealed that isolates in each clade appear to interact differently with complement proteins in human serum. In contrast to a prior report suggesting that there is no difference in serum survival between Clade 1 and Clade 2 isolates (21), we found that the Clade 2 isolates in our collection were, on average, more resistant to killing by human serum compared to Clade 1 isolates. Given the variability we observed in serum survival across all isolates, this may be simply due to testing more isolates. It could also be a phenomenon specific to our collection of isolates, or to the specific serum that we used. Consistent with prior studies demonstrating the importance of capsule in protection against serum-mediated killing (41, 42), we found that deletion of the capsule locus rendered strains of both KP ST258 clades much more susceptible to serum killing. Surprisingly, we found that many Clade 2 isolates had high serum resistance despite increased C3 binding. This phenomenon is puzzling, but has been previously described. For example, one study found that a serum-resistant KP ST258 isolate had increased C3b binding to its surface compared to serum-resistant hypervirulent KP isolates that were also tested (43). A prior study at our center also reported a KP ST258 isolate that, over the course of infection in a patient, evolved to resist serum-mediated killing despite high C3 binding (44). Our findings show that the Clade 2 capsule, perhaps due to its composition and/or structure, seems to be uniquely able to tolerate C3 binding to aid bacteria in resisting serum-mediated killing. Interestingly, clade-specific differences in C3 binding were still apparent among the capsule knockout strains, suggesting that there may be additional differences between Clade 1 and Clade 2 that contribute to increased C3 binding independent of susceptibility to human serum. In addition to increased C3 binding to Clade 2 isolates, we also observed increased C5a generation compared with Clade 1 isolates. Excess complement activation, and in particular elevated C5a levels, have been implicated in tissue damage and acute lung injury during viral infection (45, 46). In fact, several therapeutics targeting C5 and C5a generation for the treatment of COVID-19 are being developed (47–49). Our findings suggest that C5a production could be explored as a potential immunotherapeutic target for improved treatment of multidrug-resistant KP infections.

We found that a representative Clade 2 isolate had more dissemination than a representative Clade 1 isolate in a mouse model of pneumonia, and that this dissemination was likely capsule-dependent. However, we found that increased dissemination could not be explained by the high serum resistance we observed among Clade 2 isolates, since mouse serum was unable to kill any of the isolates that we tested. Most notably, mouse serum was unable to kill the capsule knockout strains, which were both highly susceptible to killing by human serum. The inability of mouse serum to kill KP isolates has been reported previously (50), and is likely due to the fact that mouse serum has very low levels of complement activity (51). It is also possible that cytolytic activity is not a primary function of murine complement, especially given that mouse serum contains a factor that inhibits binding of the terminal complement complex to target cells (52). Despite the difference in killing activity between human and mouse serum, we nonetheless found that the representative Clade 1 isolate we tested was phagocytosed more by murine macrophages than the representative Clade 2 isolate, suggesting that in mice, macrophages might play a more critical role in bacterial clearance than the complement system. Complement could instead have other functions in mice; for example, it was recently shown that C3 and C4 contribute to containment of KP during gastrointestinal colonization in mice (53).

This study had several limitations. First, the isolates we studied were all derived from a single geographic area, and some were highly genetically related. Second, we were only able to phenotype a subset of the isolates in our original clinical isolate set, and this subset contained more Clade 2 isolates than Clade 1 isolates. Additionally, we used a single representative isolate from each clade in our generation of capsule knockout strains and mouse experiments. While we demonstrated that the capsule is critical for human serum resistance and dissemination of a Clade 2 isolate in mice, we cannot definitively show that its virulence is due specifically to the Clade 2 capsule as opposed to the Clade 1 capsule. The capsule knockout strains we developed, however, are well positioned for complementation with the opposing capsule locus, and this is an important area of further investigation. Lastly, we found that Clade 2 isolates were generally able to resist serum-mediated killing despite high C3 protein binding, however the molecular mechanism(s) underlying this phenomenon remain undefined.

In summary, this study provides a systematic comparison of KP ST258 Clade 1 and Clade 2 isolates using clinical, genomic, *in vitro*, and *in vivo* strategies. We found that Clade 2 isolates were more prevalent clinically and that Clade 2-infected patients were more acutely ill than Clade 1-infected patients. We also showed that compared to Clade 1 isolates, Clade 2 isolates were more resistant to human serum and more able to disseminate to other organs in a mouse model of pneumonia. Furthermore, we identified important differences in the host response to KP ST258 infection between humans and mice that underscore the limitations of mouse models in studying complement-mediated killing of pathogens. Lastly, the increased human complement activation that we identified in Clade 2 highlights the potential for novel therapeutic strategies targeting excess C5a production. Overall, these findings demonstrate the increased virulence potential of Clade 2 and identify possible opportunities for future development of complement-targeting immunotherapeutics for KP infection.

## METHODS

### Isolate collection

A total of 172 KP ST258 isolates were collected between 2015 and 2024 from patients at the University of Pittsburgh Medical Center (UPMC). Isolates were collected as part of a genomic surveillance program (Enhanced Detection System for Healthcare-Associated Transmission, EDS-HAT) and as part of routine collection of carbapenem-resistant KP at UPMC (54, 55). This study was approved by the Institutional Review Board at the University of Pittsburgh (STUDY21040126 and STUDY22070065). Isolates were identified as ST258 through multiplex PCR (56, 57), or from the genome sequence using Kleborate version 3.2.4 (56). Clade was assigned using *cps-1* and *cps-2* primers (58), or by isolate placement on the phylogenetic tree (**Fig S2A).**

### Clinical review of patients infected with KP ST258

Review of electronic medical records was used to obtain clinical information including: age, sex, survival starting from date of isolate collection, Charlson Comorbidity Index (27), and Pitt Bacteremia Scores (28, 29, 59). For the purposes of calculating the Pitt Bacteremia Score, patients that were sedated were assigned 2 points for mental status (stuporous) and patients that were nonresponsive without sedation were assigned 4 points for mental status (comatose).

### Whole-genome sequencing and bioinformatic analysis

Whole genome sequencing was performed on the Illumina platform (NextSeq550, NextSeq1000, and NovaSeq X Plus) and accession numbers are listed in Table S4. Genome assemblies were used that had <500 contigs and did not have any QC flags when run through Kleborate v3.2.4 (56). Genomes were assembled using Unicycler v0.5.1 and annotated using Prokka v1.14.5 (60). GFF files generated through Prokka were used by Roary v3.13.0 to build a core genome alignment (61). A phylogenetic tree was constructed using the GTR GAMMA model of RAxML HPC v8.2.12 with 100 bootstraps and was visualized in iTol v7.4.2 (62, 63). Genetic relatedness of isolates was measured using Split Kmer Analysis v1.0 (SKA) to identify single nucleotide polymorphisms (SNPs) in pairwise genome-wide comparisons (64). Clusters of genetically related isolates were identified with a 10-SNP cut off. The number of plasmid replicons in each isolate genome was identified using ABRicate v0.7 to compare with the PlasmidFinder database (65, 66) with >50% nucleotide identity and >90% coverage as cutoffs. The number of resistance genes and siderophores in each isolate genome was identified using Kleborate (56).

### Capsule isolation and purification for carbohydrate composition analysis

Capsule isolation and purification was performed as previously described (67, 68). In brief, colonies were scraped from agar plates and crude capsular polysaccharide extracts were obtained via acetone precipitation. These precipitates were dialyzed and lyophilized before treatment with RNase, DNase, and proteinase. Purified and treated capsular polysaccharides were then dialyzed and lyophilized again. The lyophilized capsule polysaccharide was then sent to the Complex Carbohydrate Research Center at the University of Georgia for composition analysis.

### Serum survival assay

Overnight cultures of bacterial isolates were diluted 1:100 in LB media and subcultured at 37°C for 2.5 hours. Bacteria were pelleted by centrifugation and normalized to an OD600nm of 0.2 in 1xPBS. Four microliters of normalized bacteria were then mixed with 2µL LB media and 34 µL human or mouse serum (Complement Technology) and incubated for 30 minutes at 37°C in a sealed half-area 96-well plate. After incubation, samples were serially diluted in PBS and plated onto LB agar to obtain colony counts. Each isolate was tested with at least two biological replicates.

### Generation of capsule knockout strains using Lambda Red recombineering

KLP00155 and KLP00157 were chosen as Clade 1 and Clade 2 representatives, respectively, because they were both hybrid assembled and used as representative isolates in a prior publication (19). A lambda red machinery plasmid (pMQ763) was introduced into the target isolates (KLP00155 and KLP00157) using conjugation. This plasmid is a hygromycin-resistant variant of pMQ538 (69). Tetracycline resistance cassettes specific to each target isolate were generated using PCR primers that added 40bp flanks to each end of the cassette. The tetracycline resistance cassette was derived from plasmid pMQ809, which has *tetRA*, (unpublished) and the primers used are listed in Table S3. These flanks were homologous to the regions immediately upstream and downstream of the capsule locus of each isolate. In both target isolates, the deleted region spans from gene locus tag KLP00155_04906 (Clade 1 representative) or KLP00157_01733 (Clade 2 representative) to *uge.* The deleted regions were 28,291 bp in KLP00155 and 19,389 bp in KLP00157. Target isolates containing lambda red machinery were made electrocompetent and the lambda red machinery was induced using arabinose. The purified isolate-specific tetracycline resistance cassettes with homologous ends were then electroporated into the competent, lambda red-induced target isolates. Replacement of the capsule locus with the tetracycline cassette was verified by PCR and whole genome sequencing.

### Growth assays

Overnight cultures were normalized to an OD600nm of 0.05 and then diluted 1:5,000 in LB media. Diluted cultures were plated in a 96-well plate for growth measurement over 20 hours in a BioTek Synergy plate reader at 37°C with orbital shaking and OD600nm readings every 15 minutes.

### C3 binding ELISA

Overnight cultures were diluted 1:100 in LB media and then and subcultured for 2.5 hours at 37°C. All cultures were then normalized to an OD600nm of 0.2 in PBS and 100ul of the normalized culture was plated in a high-binding 96-well plate. The plate was then dried at 37°C overnight. The plate was then blocked with 1% bovine serum albumin in PBS for 1 hour. Blocking solution was removed and 5% normal human serum (Complement Technology) in PBS was added to each well. After a 1-hour incubation at room temperature, the plate was washed 3 times with 0.05% Tween20 in PBS and then primary antibody (1:200,000 goat anti-human C3 antibody, Complement Technology) was added to the plate. After a 1-hour incubation, the plate was washed 3 times and secondary antibody was added to the plate (1:5000 rabbit anti-goat IgG HRP-conjugated antibody, R&D Systems). After 1-hour incubation, the plate was washed 3 times and TMB substrate was added to the plate. After a 10-minute incubation, the reaction was stopped with 2N H_2_SO_4_. The absorbance was then read in a BioTek Synergy plate reader at 450nm and 540nm. At least 2 biological replicates were tested per isolate and each biological replicate had at least 5 technical replicates.

### Uronic Acid Quantification Assay

Uronic acid content of isolates was quantified using previously described methods (70). In brief, capsule was extracted using 1% zwittergent 3-14 and precipitated with ethanol. Precipitated capsule was resuspended in ultrapure water. Twenty microliters of glucuronolactone standards and samples were transferred to a 96-well plate with 120ul of sodium tetraborate in sulfuric acid and boiled. After cooling, 100ul of each sample and standard were transferred to a half-area 96-well plate for absorbance measurement at 520nm. Two microliters of 3-phenylphenol were then added to each well. After thorough mixing, the plate was measured again at 520nm. Uronic acid concentration was calculated by subtracting the second Abs520nm reading from the first reading and then calculating concentration based on the standard curve generated from the glucuronolactone standards.

### C5a ELISA

Overnight cultures were diluted 1:100 in LB media and then subcultured for 2.5 hours at 37°C before normalizing to an OD600nm of 0.2 in PBS. Normalized cultures were then incubated in 5% normal human serum (Complement Technology) in PBS for 30 minutes rotating at 37°C. These reactions were then centrifuged at 10,000xg for 5 minutes at 4°C. The supernatant was then removed and used for C5a quantification. C5a quantification was performed using the Human Complement Component C5a DuoSet ELISA from R&D systems. At least 2 biological replicates were performed for each isolate, each with at least two technical replicates.

### Mouse model of pneumonia via oropharyngeal aspiration

All animal studies were conducted with approval from the University of Pittsburgh Institutional Animal Care and Use Committee (Protocol 23073501) and conducted in compliance with its guidelines. Inoculums for infection were prepared by diluting overnight cultures 1:100 in LB media and subculturing for 2.5 hours at 37°C. The subcultures were then normalized to an OD600nm of 0.2 in PBS for wildtype isolates or 0.4 for KLP00157Δcps. The normalized inoculum was serially diluted and plated to quantify CFUs. Mice were 8-16 week old, age-matched, female, C75BL/6J mice purchased from The Jackson Laboratory. At least eight mice per group were infected. Mice were sedated with inhaled isoflurane and infected via oropharyngeal aspiration with 50ul of normalized bacterial inoculum. Bronchoalveolar lavage fluid (BALF) and tissues were harvested at 6 or 24 hours post-infection. Mice were euthanized at 6 or 24 hours via pentobarbital injection followed by cervical dislocation. To measure bacterial burden and dissemination, the upper right lung lobe, liver, and spleen were each homogenized in 1mL (2mL for liver) PBS, serially diluted in PBS, and plated on LB agar for CFU enumeration. BALF was obtained by cannulation and lavage with 1mL of PBS. BALF was centrifuged (2,000xg, 5 min) and supernatant was separated from the pelleted cells. Protein content in the BALF supernatant was measured using the Pierce BCA protein assay kit (ThermoFisher). Cells from BALF were treated with ACK lysis buffer (Gibco) to remove red blood cells and resuspended in 500ul PBS. Cells were counted by a TC20 automated cell counter (BioRad). A total of 200ul of the resuspended cells were placed on a slide using a cytospin centrifuge and the cells were stained with Hema 3 Solution II staining solutions (FisherScientific) to enable enumeration of monocytes, polymorphonuclear cells, eosinophils, and lymphocytes. For cytokine measurements, RNA from the middle right lobe, which had been snap frozen in liquid nitrogen and stored at -80°C, was extracted using the Qiagen RNeasy Mini Kit. cDNA was synthesized using the iScript cDNA synthesis kit (BioRad) and qRT-PCR was performed using Sso Advanced Universal Probe Supermix (BioRad) and target-specific FAM TaqMan real-time PCR assay primer probes (ThermoFisher). Transcript levels of each target gene were compared to those of the *HPRT* gene in PBS mock infected mice.

### Gentamicin protection assay

Immortalized bone marrow-derived murine macrophages (NR-9456 BEI Resources) were incubated with live KP isolates at a multiplicity of infection (MOI) of 10 in DMEM at 37°C for 2 hours. Gentamicin was then added at a final concentration of 1mg/mL and the macrophages with bacteria were incubated for an additional hour on ice. Macrophages were washed with PBS and then lysed with 0.2% triton X-100. The lysate was then diluted and plated to obtain colony counts. Six biological replicates were performed with at least 2 technical replicates each.

### Statistical analysis

All statistical calculations were performed in GraphPad Prism v10.6.1. Survival curves were assessed using log-rank tests. Categorical comparisons (sex of patients infected with isolates, isolate source distribution, clustering of isolates, and percentage of isolates containing siderophores and antibiotic resistance genes) were assessed using Fisher’s exact tests. Comparisons between age, Charlson Comorbidity Index, Pitt Bacteremia Score, and hospitalization lengths of patients infected with isolates were assessed using Mann-Whitney tests. Comparisons between clades or isolates in genome size, number of plasmid replicons, number of resistance genes, survival in serum, C3 binding, C5a production, BALF protein content, BALF total cell count, BALF differential cell counts, cytokine expression, bacterial burden in organ homogenates, and macrophage uptake of bacteria were also assessed using Mann-Whitney tests. Comparisons between clades or isolates of uronic acid quantity were assessed using Welch’s t-tests. Relationship between survival in human serum and C3 binding, survival in human serum and uronic acid quantity were assessed via Spearman correlation. Results were considered significant if the P-value was less than or equal to 0.05.

## ACKNOWLEDGEMENTS

We thank Robert Shanks for plasmids, advice, and help with the generation of capsule knockout strains. We thank Ellen Kline for helping process and sequence clinical KP isolates for use in this study. We thank Emma Mills for her expertise and technical assistance with comparative genomic analysis. We thank Shekina Gonzalez-Ferrer, Renata DiDonato, Hayley Ramirez, and Caitlin Preusser for their assistance with mouse experiments. We thank Laura Mike and her laboratory for providing the macrophages used and their assistance with the gentamicin protection assay. We thank the Complex Carbohydrate Research Center at the University of Georgia who analyzed the carbohydrate content of the KLP00155 and KLP00157 capsules. Their work was supported by the US Department of Energy, Office of Science, Basic Energy Sciences, Chemical Sciences, Geosciences and Biosciences Division, under award #DE-SC0015662. This work was supported by the Department of Medicine at the University of Pittsburgh, AHA predoctoral fellowship 25PRE1375562 and NIH T32 AI138954 (to NC), and NIH R01 HL107380 and HL167449 (to JFA).

## DATA AVAILABILITY

Raw Illumina sequencing reads are uploaded to NCBI Sequence Read Archive (SRA) and GenBank. BioProject numbers and BioSample numbers are listed in Table S4. Raw data associated with each isolate in this study is available in Table S4.

## SUPPLEMENTAL FIGURES

**Fig S1. Clinical features of patients infected with Clade 1 (n = 46) or Clade 2 (n = 126) KP ST258 isolates.** On the day the clinical isolate was collected, we assessed: (A) Patient age, (B) Charlson Comorbidity Index, (C) Pitt Bacteremia Score (calculated for the day each isolate was obtained and the 48 hours prior), and (D) Length of hospitalization for each patient. P ≤ 0.05: *, P ≤ 0.01: ** by Mann-Whitney test.

**Fig S2. Genetic relatedness and genomic and clinical features of KP ST258 Clade 1 (n = 25) and Clade 2 (n = 108) isolates collected from UPMC.** The midpoint-rooted phylogenetic tree was constructed using RAxML based on a core genome alignment of 4236 genes identified by Roary. K Locus, O Locus, isolate source, and year of isolation are colored as indicated. K Locus and O Locus alleles were assigned using Kleborate.

**Fig S3. Genetic relatedness and antibiotic resistance genes encoded by Clade 1 and Clade 2 isolates.** (A) Pairwise genome-wide comparisons were performed using split kmer analysis. (B) Number of plasmid replicons measured by ABRicate using the PlasmidFinder database. (C) Number of resistance genes determined by Kleborate. (A-C) P ≤ 0.01: **, P ≤ 0.0001: **** by Mann-Whitney test. (D) Percent of Clade 1 or Clade 2 isolates that have at least one gene encoding resistance to the macrolide, phenicol, sulfonamide, tetracycline, or trimethoprim drug classes or genes encoding a non-SHV beta-lactamase, SHV beta-lactamase, ESBL beta-lactamase, or carbapenemase as identified by Kleborate. P ≤ 0.05: *, P ≤ 0.0001: **** by Fisher’s exact test.

**Fig S4. Characterization of capsule knockout strains.** (A) Uronic acid quantification of wildtype (KLP00155 and KLP00157) and capsule knockout strains (KLP00155Δcps and KLP00157Δcps). Three biological replicates were performed per isolate. P ≤ 0.01: ** by Welch’s t-test. (B and C) Bacterial isolates were normalized to an OD600 of 0.05 and then diluted 1:500 in LB media. Isolates were grown for 20 hours at 37°C with orbital shaking and OD600 measurements were collected every 15 minutes. Three biological replicates (with four technical replicates each) were performed per isolate.

**Fig S5. Correlation between survival in human serum and either C3 binding or uronic acid.** Association of (A) C3 binding and serum survival, and (B) Uronic acid quantity and serum survival of Clade 1 isolates. Associations were assessed using a Spearman correlation.

**Fig S6. Lung inflammation after infection with Clade 1 or Clade 2 isolates.** Bronchoalveolar lavage fluid (BALF) was collected from mice 6- and 24-hours post infection (HPI). For 6 HPI: KLP00155 n=8 and KLP00157 n=6. For 24 HPI: KLP00155 n=8 and KLP00157 n=7. (A) Protein in BALF was quantified using a BCA assay. (B) Total cell count in BALF was quantified using an automated cell counter. (C) Cells from BALF at 6 and 24 HPI were concentrated on a microscope slide and stained for differential cell counting of monocytes (MONO), polymorphonuclear cells (PMN), eosinophiles (EOS), and lymphocytes (LYM). (D and E) Cytokine levels were measured using TaqMan RT-qPCR on RNA isolated from the middle right lobe at (D) 6 HPI (KLP00155 N=8, KLP00157=7) and (E) 24 HPI (KLP00155 and KLP00157 N=8) and compared to *HPRT* gene expression levels of PBS mock-infected mice. P-value was calculated by Mann-Whitney test.

