## Supplemental Figures for "Differential virulence potential of different clades of multidrug-resistant *Klebsiella pneumoniae* ST258"

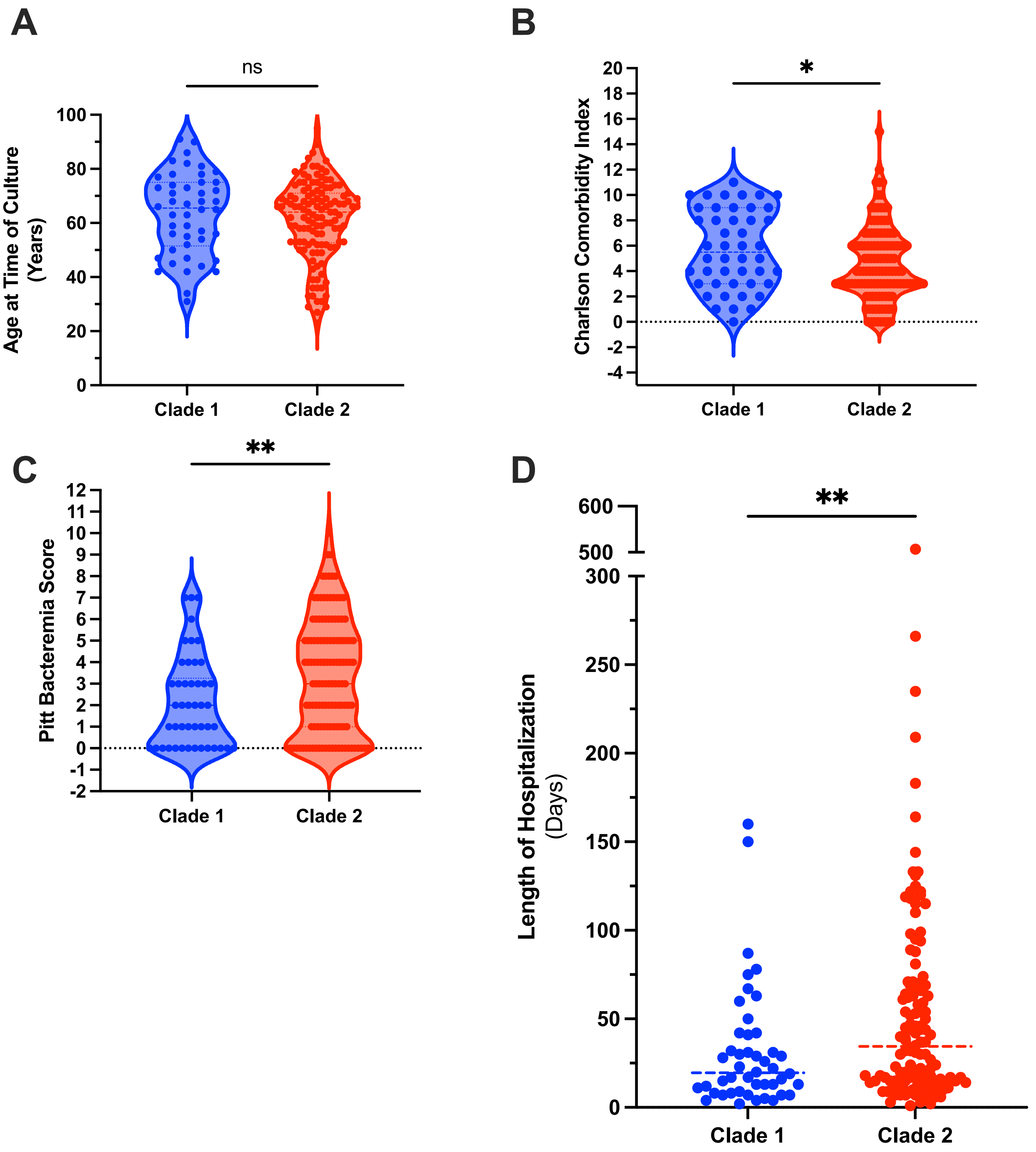


**Fig S1. Clinical features of patients infected with Clade 1 (n = 46) or Clade 2 (n = 126) KP ST258 isolates.** On the day the clinical isolate was collected, we assessed: (A) Patient age, (B) Charlson Comorbidity Index, (C) Pitt Bacteremia Score (calculated for the day each isolate was obtained and the 48 hours prior), and (D) Length of hospitalization for each patient. $P \leq 0.05$: *, $P \leq 0.01$: ** by Mann-Whitney test.

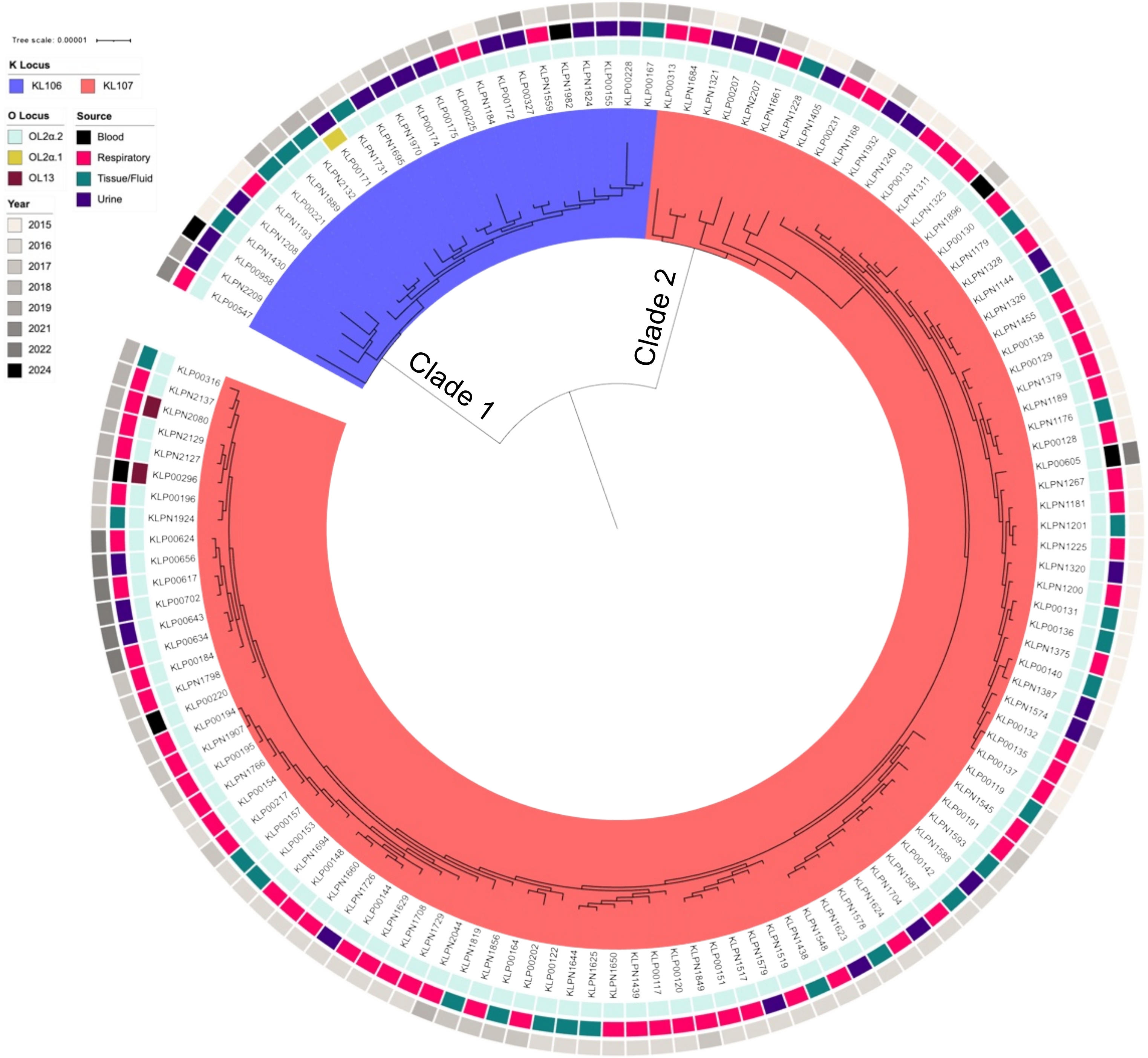


**Fig S2. Genetic relatedness and genomic and clinical features of KP ST258 Clade 1 (n = 25) and Clade 2 (n = 108) isolates collected from UPMC.** The midpoint-rooted phylogenetic tree was constructed using RAxML based on a core genome alignment of 4236 genes identified by Roary. K Locus, O Locus, isolate source, and year of isolation are colored as indicated. K Locus and O Locus alleles were assigned using Kleborate.

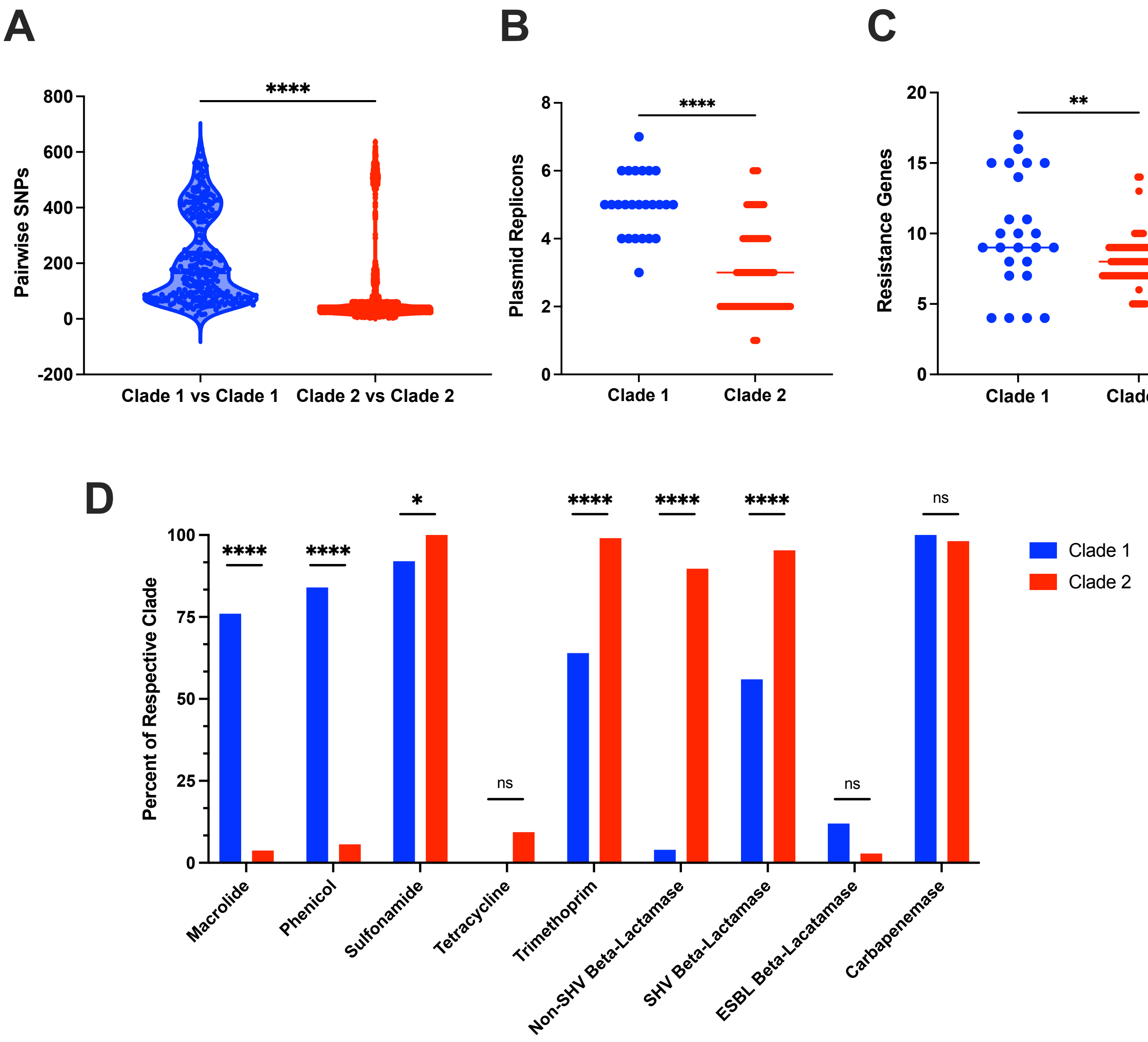


**Fig S3. Genetic relatedness and antibiotic resistance genes encoded by Clade 1 and Clade 2 isolates.** (A) Pairwise genome-wide comparisons were performed using split kmer analysis. (B) Number of plasmid replicons measured by ABRicate using the PlasmidFinder database. (C) Number of resistance genes determined by Kleborate. (A-C) $P \leq 0.01$: **, $P \leq 0.0001$: **** by Mann-Whitney test. (D) Percent of Clade 1 or Clade 2 isolates that have at least one gene encoding resistance to the macrolide, phenicol, sulfonamide, tetracycline, or trimethoprim drug classes or genes encoding a non-SHV beta-lactamase, SHV beta-lactamase, ESBL beta-lactamase, or carbapenemase as identified by Kleborate. $P \leq 0.05$: *, $P \leq 0.0001$: **** by Fisher's exact test.

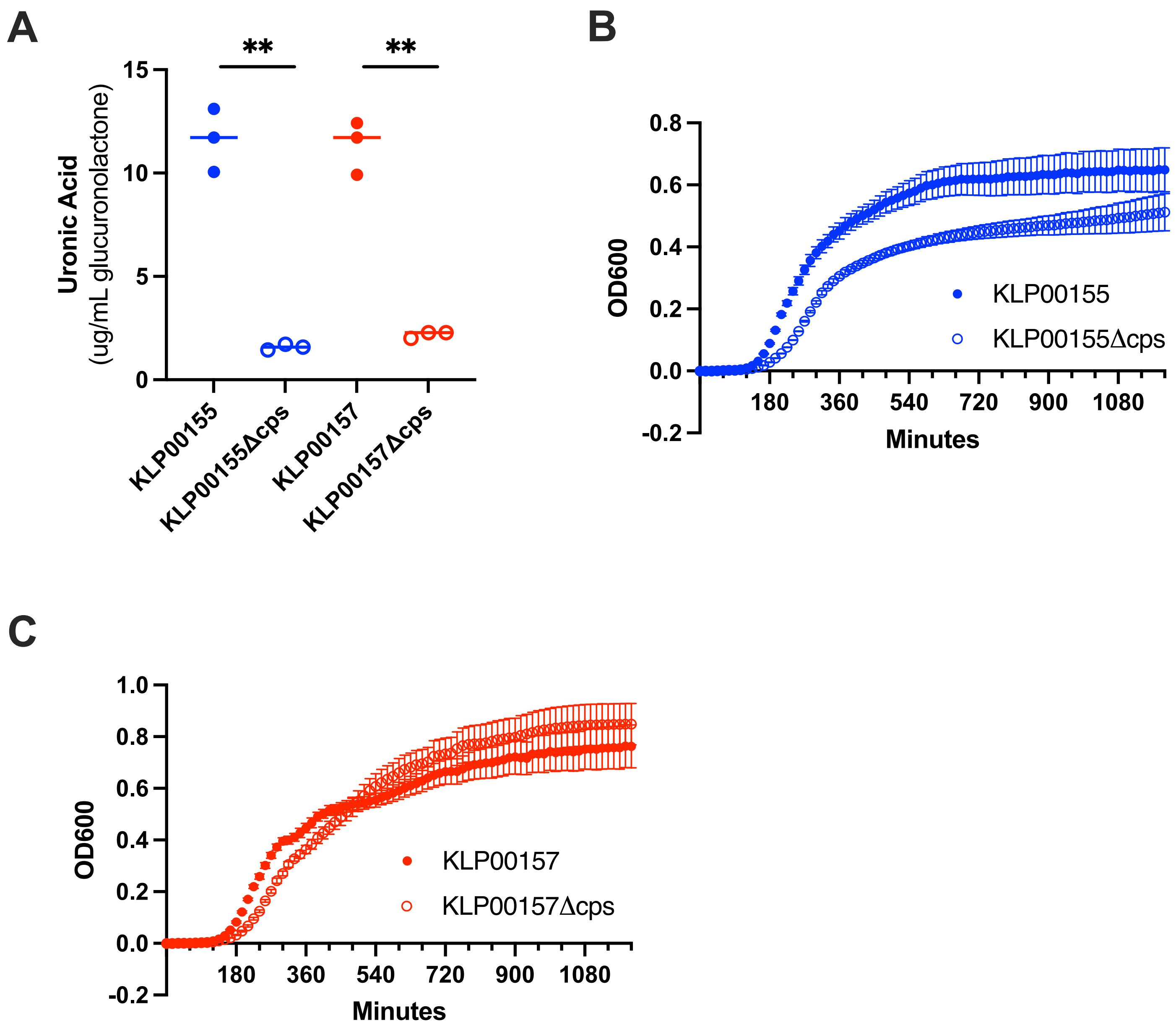


**Fig S4. Characterization of capsule knockout strains.** (A) Uronic acid quantification of wildtype (KLP00155 and KLP00157) and capsule knockout strains (KLP00155Δcps and KLP00157Δcps). Three biological replicates were performed per isolate. $P \leq 0.01$: ** by Welch's t-test. (B and C) Bacterial isolates were normalized to an OD600 of 0.05 and then diluted 1:500 in LB media. Isolates were grown for 20 hours at 37°C with orbital shaking and OD600 measurements were collected every 15 minutes. Three biological replicates (with four technical replicates each) were performed per isolate.

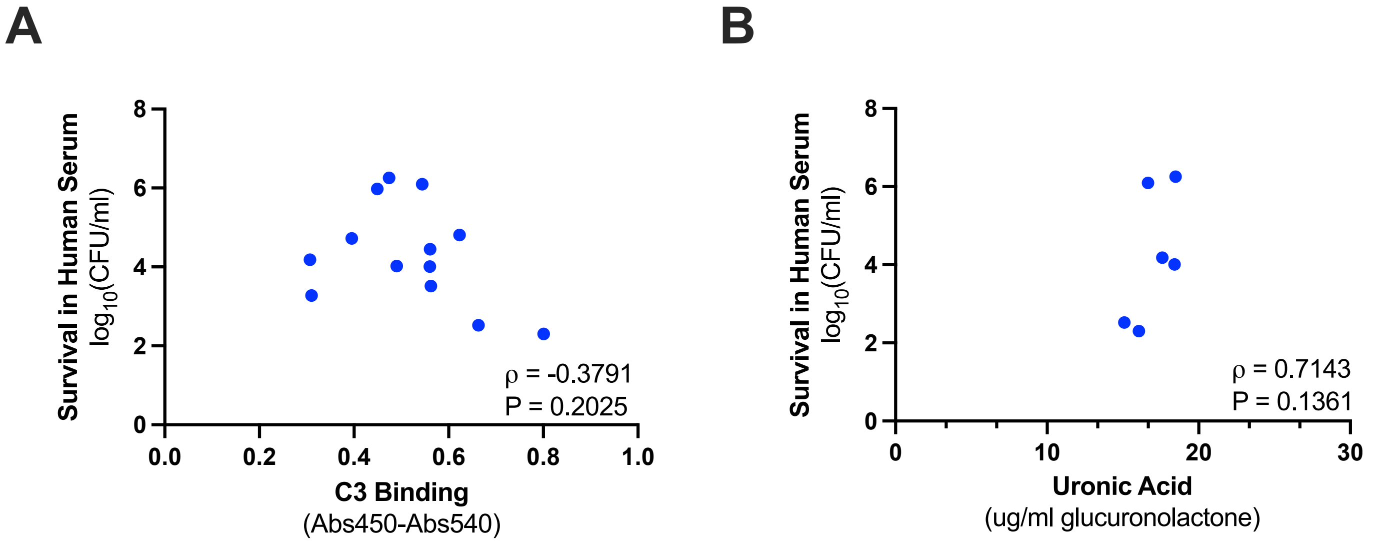


**Fig S5. Correlation between survival in human serum and either C3 binding or uronic acid.** Association of (A) C3 binding and serum survival, and (B) Uronic acid quantity and serum survival of Clade 1 isolates. Associations were assessed using a Spearman correlation.

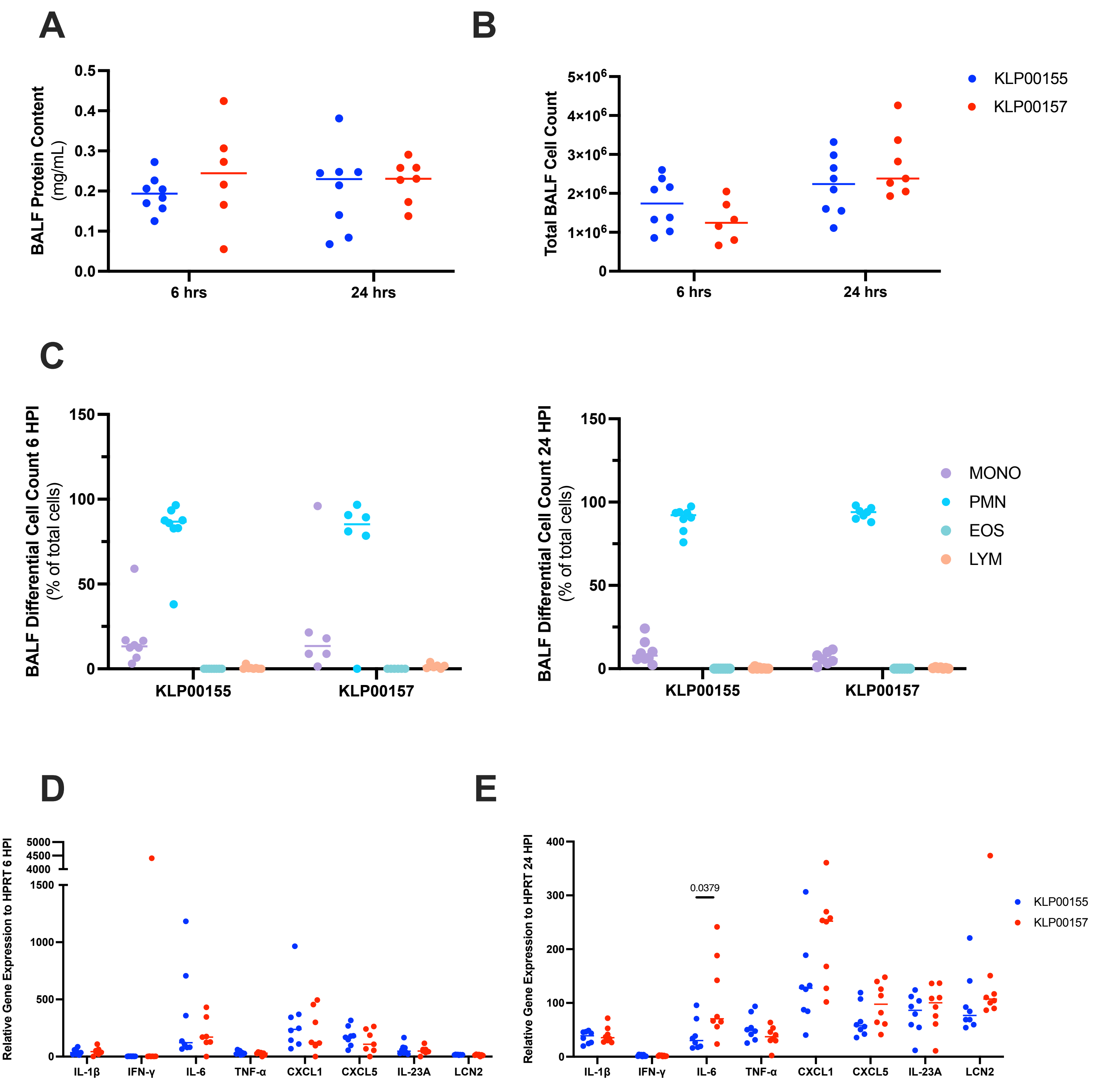


**Fig S6. Lung inflammation after infection with Clade 1 or Clade 2 isolates.** Bronchoalveolar lavage fluid (BALF) was collected from mice 6- and 24-hours post infection (HPI). For 6 HPI: KLP00155 n=8 and KLP00157 n=6. For 24 HPI: KLP00155 n=8 and KLP00157 n=7. (A) Protein in BALF was quantified using a BCA assay. (B) Total cell count in BALF was quantified using an automated cell counter. (C) Cells from BALF at 6 and 24 HPI were concentrated on a microscope slide and stained for differential cell counting of monocytes (MONO), polymorphonuclear cells (PMN), eosinophiles (EOS), and lymphocytes (LYM). (D and E) Cytokine levels were measured using TaqMan RT-qPCR on RNA isolated from the middle right lobe at (D) 6 HPI (KLP00155 N=8, KLP00157=7) and (E) 24 HPI (KLP00155 and KLP00157 N=8) and compared to *HPRT* gene expression levels of PBS mock-infected mice. P-value was calculated by Mann-Whitney test.
